# ChatGPT as a medical doctor? A diagnostic accuracy study on common and rare diseases

**DOI:** 10.1101/2023.04.20.23288859

**Authors:** Lars Mehnen, Stefanie Gruarin, Mina Vasileva, Bernhard Knapp

## Abstract

Seeking medical advice online has become popular in the recent past. Therefore a growing number of people might ask the recently hyped ChatGPT for medical information regarding their conditions, symptoms and differential diagnosis. In this paper we tested ChatGPT for its diagnostic accuracy on a total of 50 clinical case vignettes including 10 rare case presentations. We found that ChatGPT 4 solves all common cases within 2 suggested diagnoses. For rare disease conditions ChatGPT 4 needs 8 or more suggestions to solve 90% of all cases. The performance of ChatGPT 3.5 is consistently lower than the performance of ChatGPT 4. We also compared the performance between ChatGPT and human medical doctors. We conclude that ChatGPT might be a good tool to assist human medical doctors in diagnosing difficult cases, but despite the good diagnostic accuracy, ChatGPT should be used with caution by non-professionals.

## Introduction

A large number of people seek medical advice online and most have tried to “google” for symptoms they are experiencing. Therefore, a wide range of online possibilities for self-diagnosis exist. These include classical search engines and governmental webpages such as the National Health Service (NHS) UK as well as specifically developed websites and mobile phone apps often referred to as symptom checkers (see e.g. [1]).

Recently, transformer models [2], large language models (LLMs) [3] and especially Generative Pre-trained Transformer (GPT) models available as chatbot (e.g. chatGPT) [4] have gained considerable attention in the scientific community and wider public. These GPT models depart from previous LLMs in terms of number of parameters, training set size and supervised/reinforcement fine-tuning approaches [3,5].

ChatGPT is a general purpose language model that was, in contrast to classical symptom checkers, not developed specifically to solve clinical case vignettes. However, ChatGPT has the potential to semantically understand user input and provide answers for a wide variety of tasks ranging from asking general knowledge questions, programming and mathematics to scientifically (sounding) text-generation and law.

In the current study we tested the ability of ChatGPT to find the correct diagnoses for full-text clinical case vignettes of patients. This includes common disease presentations that most human medical doctors should relatively easily solve as well as more difficult rare disease presentations that are less common [6].

## Methods

### Clinical case vignettes

To test the diagnostic accuracy of ChatGPT we used a total of 50 clinical case vignettes of which 40 were common complaints and 10 rare diseases. For common complaints, we used 40 of the 45 case vignettes collected by [1]. We excluded 5 cases which contain the correct diagnosis as a symptom within the vignette text e.g. a case vignette where a patient complains about vomiting with the correct diagnosis “vomiting” is not sufficiently challenging. The 10 rare disease cases were generated by randomly choosing rare disorders with an orphan drug with positive status from the European Medicines Agency (EMA, [7]). The rare disease name was used as a query on https://pubmed.ncbi.nlm.nih.gov/ and the case description of the first matching article was used.

All 50 clinical case vignettes including the correct diagnoses are available from the supplementary material.

### Use of ChatGPT

ChatGPT was prompted with the phrase “What are the 10 most likely diagnoses for this patient?” followed by the clinical case vignette as full text. No symptom extraction, as necessary for most symptom checkers (see e.g. [1,8]), was performed.

The output of LLMs including ChatGPT is not deterministic therefore we prompted each vignette 3 times in independent chats. ChatGPT was used in version 3.5 as well as in version 4. This totals 300 (=50×3×2) prompts and 3000 suggested diagnoses.

### Human medical doctors

We also presented the 50 case vignettes to 3 different human medical doctors (MDs). The MDs were instructed to suggest a minimum of 1 and a maximum of 10 potential diagnoses per case vignette where the first suggestion is the most likely diagnosis, the second suggestion the second most likely diagnosis etc. The MDs were also instructed not to use a search engine. Anonymity was guaranteed to all 3 participating MDs.

### Assessment of correct answers

All suggested diagnoses were compared by another human medical doctor with the correct diagnosis provided by the respective case vignette. A case was considered correct if either a direct match (e.g. “acute otitis media” vs “acute otitis media”) or a direct hierarchical relation with the correct diagnosis (e.g. “acute pharyngitis” vs “pharyngitis”, “GM2 gangliosidosis” vs Tay–Sachs disease”, “stroke” vs “ischemic stroke” etc.) existed.

### Presentation of diagnostic accuracy

We expressed the correctness of suggested diagnoses as topX accuracy i.e. how many percent of cases are solved using a maximum of X suggested diagnosis. For example a top1 accuracy of 100% would mean that all clinical case vignettes were solved by the first suggested diagnosis. If 7 out of 10 cases would be solved by the first suggested diagnosis and one additional case by the second suggested diagnosis then the top1 accuracy would be 70% and the top2 accuracy would be 80%.

For the diagnoses agreement hypothesis-test between ChatGPT 3.5, ChatGPT 4 and the correct diagnosis, we used Fleiss’ kappa [9], which is an extension of Scott’s pi statistic [10]. This choice was made as Scott’s pi and Cohen’s kappa are limited to two sets of diagnoses, Fleiss’ kappa can accommodate any number of sets of diagnoses assigning categorical ratings.

## Results

### Diagnostic accuracy on common cases

In Figure 1 the ChatGPT 3.5 and 4 results are shown. Within 2 suggested diagnoses (top2 accuracy) more than 90% of all cases were solved by ChatGPT 3.5. The results for ChatGPT 4 are even superior and 100% of all cases were solved in all 3 prompt versions within 3 suggested diagnoses (top3 accuracy). The results of ChatGPT 4 are statistically significantly better than the results of ChatGPT 3 (Wilcoxon signed rank test,alpha=0.05, p<0.007).

**Figure 1:**
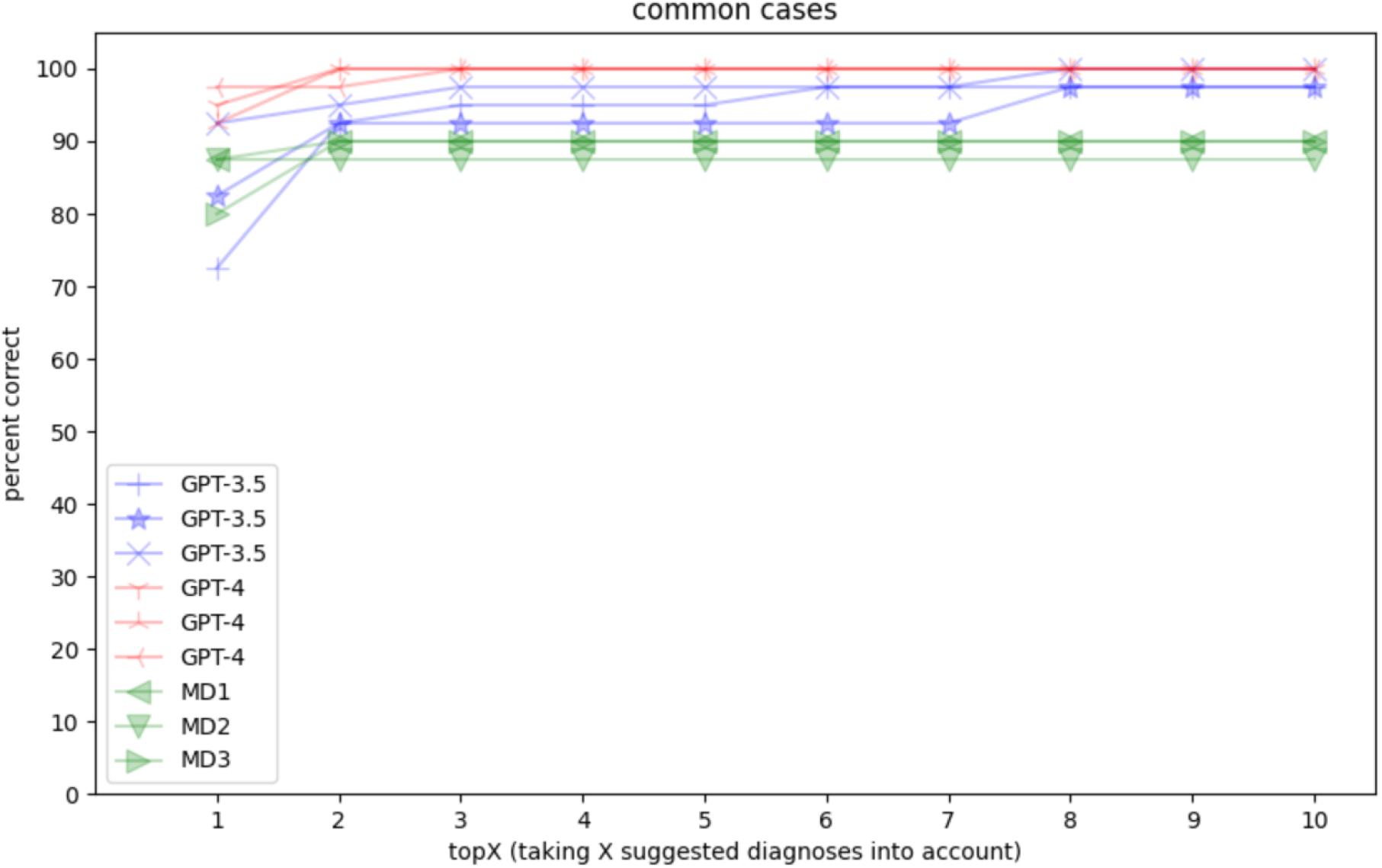
Diagnostic accuracy of ChatGPT and 3 MDs on 40 common case clinical vignettes allowing a maximum of 10 diagnosis suggestions.

A Fleiss diagnosis agreement hypothesis-test for the first GPT returned results against the given correct diagnoses yielded substantial agreement (p<0.0001). This means that the diagnoses suggested by chatGPT are significantly similar to the correct diagnoses.

The MDs solved around 90% of all cases within 2 suggestions but none of them reached 100%. For details we refer to the discussion section.

### Diagnostic accuracy on rare cases

Rare cases were a more challenging assignment for ChatGPT (see Figure 2). ChatGPT 3.5 reaches on average just 60% correct within 10 suggestions (top10 accuracy) and only 23% of the correct diagnoses were listed as the first result.

**Figure 2:**
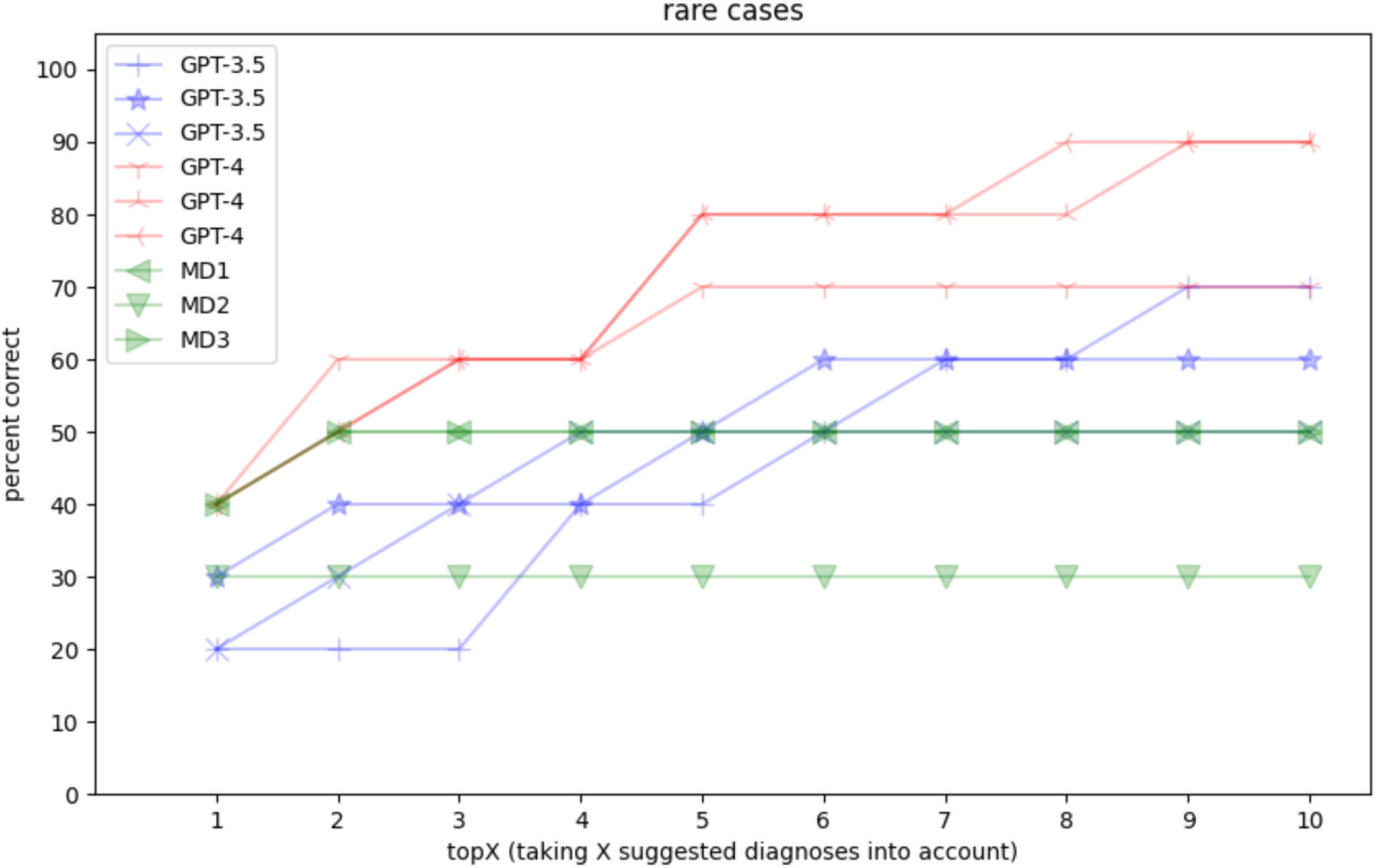
Diagnostic accuracy of ChatGPT and 3 MDs on 10 rare case vignettes allowing a maximum of 10 diagnosis suggestions.

ChatGPT 4 is more successful than ChatGPT 3.5 (Wilcoxon signed rank test, alpha=0.05, p<0.004) but still far from the performance of the common cases. 40% of all cases are solved with the first suggested diagnosis while it takes 8 or more suggestions to reach a 90% diagnostic accuracy. A performance of 100% is never reached by any of the individual GPT models. However, there is not a single case vignette that is never solved by any of the models i.e. running ChatGPT 4 three times yields 3×10 suggestions, which include the correct diagnosis for each case at least once. This means that running a model multiple times on the same input prompt can improve results slightly (here from 90% to 100%).

Also here we performed a Fleiss test giving a moderate agreement (z=9.0837, p<0.0001). This means that the diagnoses suggested by chatGPT are similar to the correct diagnoses. The observed agreement is not by chance.

The MDs solved between 30 and 40% of all cases with their first suggested diagnosis. For 2 MDs the diagnostic accuracy increases further to 50% using 2 suggested diagnoses. Also here we refer to the discussion section.

## Discussion

### ChatGPT achieves high accuracies

Our results (Figure 1) show that ChatGPT reaches quite a high accuracy when solving common clinical case vignettes. This result is in agreement with previous findings showing that ChatGPT is able to complete a major law exam in the top 10% of all humans of the year and that ChatGPT also shows human-like performance for many other academic and professional exams [5]. Also in other medical settings such as providing information for cancer patients ChatGPT has shown remarkable ability [11].

However, we found that the rare case vignettes were much more challenging for ChatGPT (Figure 2). These results need to be put in perspective as such cases are in general very challenging.

### ChatGPT does not learn case vignettes by heart but semantically understands them

As ChatGPT is likely to be trained on the whole Internet it could be argued that chatGPT learned the case vignettes by heart, or in machine learning language: test set and training set overlap. While this might partially be true there is a number of arguments against this concern:

1. The common case test set was extracted from the PDF format “data supplement” of [1]. The PDF contains the cases as one of many (in machine learning terms) poorly formatted tables starting on page 6 of 19. When ChatGPT is asked if it also indexes tables in PDFs it answers: “Tables within PDFs can be part of the training data if the text extraction process is able to successfully parse and represent them. However, the quality of the extracted text may vary, and complex formatting or poorly structured tables can lead to data loss or misinterpretation.” When further asked if it is possible to check if a specific table was used, ChatGPT answers “It is not possible to determine if a specific table was used during the training of an AI language model like GPT-4. […] The models do not retain explicit knowledge of individual sources or specific pieces of content.”. ChatGPT further suggests asking specific questions about a piece of information to determine if it was used in the model. Therefore we prompted ChatGPT with the case vignettes and asked if it could give the URL, first author or journal name. To all 3 individual questions ChatGPT answered that it does not know the answer and that using a standard search engine might be more appropriate to answer the question.
2. If a model is trained on billions of documents the contribution of a single document might not weigh very heavily on the overall model. It is also unlikely that explicit information is saved in a neural network model.
3. Being able to diagnose clinical vignettes was not the aim of the development of ChatGPT. This ability is likely a side-effect.
4. ChatGPT often does not give the exact wording of the correct answer but a medical synonym.
5. ChatGPT reasons why a certain diagnosis might be right and this reasoning is not given in the solution of the clinical case vignettes. This is further explained below.

When prompted with the following case vignette:

*“A 48-year-old woman with a history of migraine headaches presents to the emergency room with altered mentalstatus over the last several hours. She was found by her husband, earlier in the day, to be acutely disoriented and increasingly somnolent. On physical examination, she has scleral icterus, mild right upper quadrant tenderness, and asterixis. Preliminary laboratory studies are notable for a serum ALT of 6498 units/L, total bilirubin of 5.6 mg/dL, and INR of 6.8. Her husband reports that she has consistently been taking pain medications and started taking additional 500 mg acetaminophen pills several days ago for lower back pain. Further history reveals a medication list with multiple acetaminophen-containing preparations.”*

ChatGPT not only states the correct diagnosis (“Acute liver failure”) directly and indirectly in the first and second result:

*“1. Acute liver failure (ALF): The elevated ALT, bilirubin, INR, and history of taking multiple acetaminophen-containing medications suggest potential acetaminophen toxicity leading to ALF*.

*2. Acetaminophen overdose: The patient’s recent increased use of acetaminophen and the presence of multiple acetaminophen-containing medications on her list could indicate an unintentional overdose.”*

but also justifies why it comes to this suggestion including the mapping of numerical laboratory values. Going even further ChatGPT suggests alternative diagnoses that would explain the symptoms if additional information in the case vignette would be present e.g.

*“5. Alcoholic hepatitis: Although not mentioned in the history, alcohol use could be a potential cause of liver injury, resulting in similar symptoms and laboratory findings.”*

Taken together this suggests that ChatGPT-4 does not only copy and paste medical diagnosis from matching books/papers/webpages but actually semantically understands and reasons about case vignettes.

### Why rare diseases are important

We decided to also include 10 rare disease cases in our study. Rare diseases are defined as conditions that affect only a small number of people, typically fewer than 1 in 2000 individuals in the population [12]. About 5000 to 8000 rare diseases exist depending on source and exact definition [6]. These diseases are often chronic, debilitating, and life-threatening, and their rarity poses significant challenges to diagnosis, treatment, and research. Many rare diseases are genetic in origin, resulting from mutations in single genes or complex interactions between multiple genes and environmental factors. Rare diseases often have a profound impact on patients, their families, and society, underscoring the urgent need for better understanding and management of these conditions [13]. Hence correctly diagnosing these rare conditions can be especially interesting and we included 10 such cases into our analysis.

### Why the comparison between ChatGPT and human medical doctors is not fair

When carrying out a benchmark on diagnostic accuracy of an AI model, intuitively the question arises how human MDs would perform on the same test set. While the results are shown in Figure 1 and 2 we believe that the way of diagnosing considerably differs between MDs and an AI model as ChatGPT. These differences include:

1. MDs usually see their patients face-to-face and an appointment is an interactive dialogue with follow-up questions and not a rigid case description.
2. In face-to-face consultations, MDs can gain additional insight about their patients by observing visual cues and behaviors, such as facial expressions and body language and perform a physical examination.
3. Before a final diagnosis is stated usually further workup such as follow-up laboratory tests or imaging studies are carried out.
4. In our study clinical case vignettes were distributed over many areas of medicine. In practice, MDs who specialize in specific subfields are often consulted for complex health issues. Consequently, a single doctor is not expected to be an expert in all types of diseases.
5. MDs were instructed not to use an internet search engine. In practice MDs sometimes conduct literature research for hints in difficult cases.
6. MDs never used the possibility to give 10 differential diagnoses in our study. The mean number of suggestions was only 1.37 and the maximum was 3 (ChatGPT always gave exactly 10 as instructed). We also observed that most of the time either the first suggested diagnosis of an MDs was right or the case was not solved at all.

In summary, we think that drawing a direct comparison between ChatGPT and human MDs is not fair, and as a result, we refrain from conducting significance tests or commenting on the direct performance comparison between ChatGPT and human MDs.

### ChatGPT cannot replace a human medical doctor

While the above reported ChatGPT diagnostic accuracy results are surprisingly good, ChatGPT itself states that it cannot and shall not replace a human medical doctor. This is for example phrased like:

*“I’m not a doctor, but I can try to offer some information on possible diagnoses based on the symptoms you provided. It’s important to consult with a healthcare professional for a proper evaluation. That being said, the following conditions might be considered for this patient: [*…*]”*

and ChatGPT concludes its response with a sentence like:

*“Remember, it’s important to consult with a healthcare professional for a proper evaluation and diagnosis.”*.

On these grounds we believe that ChatGPT is a potentially mighty tool to assist in diagnosis but shall not be used without getting further advice from a human medical doctor before drawing any conclusions or starting a treatment.

## Data Availability

All data produced in the present study are available upon reasonable request to the authors

## Ethics statement

No experiments on stem cells, animals or humans were performed. No personal data was used. All used input data is publicly accessible.

